# Effects of high-intensity interval training in the treatment of women with lipedema: a randomized controlled crossover trial (The LipidEx study)

**DOI:** 10.1101/2025.01.21.25320877

**Authors:** Julie Caroline Sæther, Randi Merete Presthus, Randi Undebakke, Lise Madsen, Ina Lønnebakken Norberg, Sunniva Kwapeng, Katrine Mari Owe, Gabriele Erbacher, Tobias Bertsch, Julianne Lundanes, Siren Nymo, Guro Fanneløb Giskeødegård, Turid Follestad, Arnt Erik Tjønna, Anja Bye

**Affiliations:** Department of Circulation and Medical Imaging, Norwegian university of Science and Technology, Norway; Clinic of Cardiology, St.Olavs hospital, Trondheim, Norway; Clinic of Rehabilitation, St.Olavs hospital, Trondheim, Norway; Department of Clinical Medicine, University of Bergen, Norway; Heimdal Physiotherapy Clinic, Trondheim, Norway; Trondheim and surrounding area lymphedema and lipedema association, Trondheim, Norway; Norwegian Research Centre for Women’s Health, Oslo University Hospital, Oslo, Norway; Földi Clinic Hinterzarten – European Center of Lymphology in the Black Forest, Germany; Department of Clinical and Molecular Medicine, Faculty of Medicine and Health Sciences, Norwegian University of Science and Technology, Trondheim, Norway; Nord-Trøndelag Hospital Trust, Clinic of Surgery, Namsos Hospital, Norway; The Department of Public Health and Nursing, Faculty of Medicine and Health Sciences, Norwegian University of Science and Technology, Trondheim, Norway; Center for Obesity Research and Innovation, Clinic of Surgery, St. Olavs hospital – Trondheim University Hospital, Trondheim, Norway; Department of Clinical and Molecular Medicine, Norwegian University of Science and Technology, Trondheim, Norway; Clinical Research Unit Central Norway, St. Olavs Hospital, Trondheim Norway

## Abstract

Lipedema is a chronic adipose tissue disorder that primarily affects women. Its etiology remains unclear and involves abnormal accumulation of fat, mainly in the lower limbs. The condition leads to both physical and psychological morbidity, often with negative impact on daily life. Effective treatment options remain limited and are primarily conservative, including compression therapy and pain relief. Given that endurance training has demonstrated benefits in pain reduction and obesity management in other patient populations, the LipidEx study aims to investigate high-intensity interval training (HIIT) as a novel therapeutic approach for women with lipedema. We will conduct a crossover randomized controlled trial (RCT) to evaluate the effects of 12 weeks of HIIT compared to a control period of usual care. The primary outcome is change in pain, while secondary outcomes include changes in adipose tissue mass and quality of life.

## Introduction

Lipedema is a complex chronic adipose tissue disorder that primarily affects women^1^. The condition is characterized by abnormal and disproportional adipose tissue distribution, with excess subcutaneous fat deposits mainly in the lower extremities.^2^ Lipedema can result in significant physical and psychological morbidity, including chronic pain or discomfort, restricted mobility, psychological vulnerability and thereby impaired quality of life.^3-9^

There is limited research and knowledge on lipedema and a lack of consensus regarding etiology, diagnosis, and treatment. However, data suggest that lipedema is a hormonally driven disorder with a genetic predisposition.^10^ Hormonal changes during puberty, pregnancy, and menopause are known to exacerbate lipedema symptoms, and having a family history of lipedema, obesity, and/or a sedentary lifestyle are known aggravating risk factors.^10-13^ Nevertheless, patients are often unrecognized or misdiagnosed with obesity, which delay both diagnosis and treatment of these patients.^10,14,15^

Current treatment options for lipedema are limited and a strong evidence base is lacking. While current interventions, such as compression therapy, manual lymphatic drainage, and liposuction, may provide some symptomatic relief, they do not address the underlying pathology of lipedema.^10,16,17^ Exercise is in general proposed as a potential non-surgical treatment option, with the intention to reduce adipose tissue mass, inflammation, and pain, and to improve lymphatic and circulatory function.^10,18^ However, only a few studies have explored the effects of exercise in lipedema patients and there is currently no consistency to whether exercise improves lipedema.^19,20^ Despite the lack of knowledge, the existing treatment guidelines for lipedema promote a healthy lifestyle with weight control measures, including physical activity.^21^ For the general population, the World Health Organization (WHO) recommends a weekly effort of either 150–300 minutes of moderate intensity physical activity, 75–150 minutes of high intensity, or a combination of the two.^22^ Unfortunately, there are no specific guidelines for physical activity for individuals with chronic pain.^22,23^ Several studies have examined what types of exercise that are suitable for achieving pain relief, as well as the duration and intensity required to achieve this.^23,24^ It appears that a minimum of ten minutes of exercise is necessary to achieve a pain-relieving effect.^25-27^ At the same duration of exercise, higher intensities, as quantified by maximal oxygen uptake (VO_2max_) result in a stronger pain-relieving effect.^28,29^ Several studies have concluded that strenuous physical activity reduces the risk of sickness absence and disability to a greater extent than moderate and light physical activity.^30,31^ Furthermore, continuity in physical activity appears to be an important element for individuals with chronic pain.^32^ Potential contributors to the positive effects of continuity on pain relief may include the long-term impact of regular exercise on factors such as stress, sleep, physical function, depression, anxiety, well-being, and disease development.^33-37^ In addition, physical activity seems to be well tolerated by patient groups with chronic pain, and studies report high level of relief and compliance.^38-41^ Considering this, regular endurance training at high intensity may have the potential to improve pain levels also in lipedema patients.

In addition to the effects on chronic pain, exercise training at high intensity has previously been shown to reduce body mass and to improve body composition in obese and overweight individuals.^41-43^ Exercise training at high intensity is also known to reduce cardiovascular risk factors and improve metabolic fitness in adults with overweight and obesity.^41^ Whether these observations is transferable to patients with lipedema remains as open questions.

The etiology of lipedema is unknown, but progression is associated with chronic inflammation and abnormal immune response.^10^ In line with this, adipocyte hypertrophy with increased fibrosis and a distinct gene expression profile associated with infiltration of macrophages is demonstrated in lipedema tissue.^44^ Thus, integrating analyses of blood and fat samples into exercise-training studies for patients with lipedema can facilitate a deeper understanding of the condition’s pathophysiology and processes associated with improvements in lipedema-related symptoms. These analyses will provide insights into cellular and molecular mechanisms governing the systemic and local effects of exercise which is crucial for developing more effective treatments and management strategies for individuals affected by lipedema.

Recently, our research group was the first to examine the effects of high intensity interval training (HIIT) in women with lipedema (ongoing trial, ClinicalTrials.gov NCT05488977). This feasibility study confirmed that HIIT is well tolerated among women with lipedema, and 22 participants have completed the 8-week exercise intervention without any adverse events occurring. Furthermore, all of the participants who attended the first exercise session, also completed the rest of the study, indicating good adherence. Based on this, HIIT may represent a promising treatment strategy for women with lipedema that may improve current guidelines for lipedema management and contribute to a change in clinical practice.

To pave the way for more effective and targeted interventions, we now aim to investigate the effects of a 12-week supervised HIIT intervention on pain levels, body composition, including adipose tissue mass, and quality of life in women with lipedema (**Table 1**). We will supplement this with exploratory analyses of inflammatory markers, metabolites, and lipoprotein subfractions in blood and adipose tissue samples to gain insights on cellular changes induced by HIIT in women with lipedema.

**Table 1.** Design Table

| Question | Hypothesis | Sampling plan | Analysis Plan | Rationale for deciding the sensitivity of the test for confirming or disconfirming the hypothesis | Interpretation given different outcomes | Theory that could be shown wrong by the outcomes |
| --- | --- | --- | --- | --- | --- | --- |
| 1.Determine whether 12 weeks of HIIT improve pain levels in women with lipedema. | H1: 12 weeks of HIIT reduces pain levels in lipedema patients.<br>H0: 12 weeks of HIIT has no effect on pain levels in lipedema patients. | Based on power calculation on our recent feasibility study (ClinicalTrials.gov NCT05488977), a sample size of 58 women with lipedema will be recruited. This allows for 80% statistical power at a significance level of 0.05. | We will compare changes in self-reported pain in the intervention period and in the control period using the BPI questionnaire.<br>We will apply LMM with repeated measures (random intercept model) using age and baseline adipose tissue mass (%) in the lower limbs as covariates to compare the changes in pain levels during the intervention period as compared to the control period. Model assessment using various types of residual analyses may lead to | Limited data exists to make an accurate prediction of the expected size of the effect of HIIT on pain levels (and more) in lipedema patients. We used data from our recent feasibility study (ongoing trial, ClinicalTrials.gov NCT05488977) to calculating the number of patients needed to obtain a power of 80% at a significance level of 0.05. We used the collected data and observations from 22 available participants. The mean self-reported pain level at baseline for the lipedema patients | If we find a significant difference ($p < 0.05$), we will interpret this as evidence that regular exercise reduces pain in lipedema patients.<br>If no significant difference is found ($p < 0.05$), we will interpret this as we have no evidence that regular exercise has a large effect pain levels. | N/A |
|  |  |  | performing the LMM on a transformed scale. | were 55 points (based on RAND-36), and the standard deviation was 23. We consider a reduction in pain levels of 10 points to be clinically relevant, which equals a change to 65 points (inversed pain scale). Based on this, a sample size of 44 participants is needed to achieve a power of 80% at a significance level of 0.05 (assuming a correlation of 0.5 between pre- and post-measures of pain). When accounting for up to 30% drop out (due to the length of the trial), we would need to include 58 patients. |  |  |
| 2. Determine whether 12 weeks of HIIT reduces total adipose tissue mass and adipose tissue mass in the lower limbs of | H1: 12 weeks of HIIT will reduce total adipose mass and adipose mass in the lower limbs in lipedema patients. | N= 58, the same sample detailed above are used. | We will compare changes in adipose tissue mass (total and lower limbs) in the intervention period and the control period using | The same sample detailed above are used. | If we find a significant difference ($p < 0.05$), we will interpret this as evidence that regular exercise | N/A |
| women with lipedema. | H0: 12 weeks of HIIT have no effect on total adipose mass and adipose mass in the lower limbs in lipedema patients. | | a bioimpedance weight (InBody 770).<br>We will apply LMM with repeated measures (random intercept model) using age as covariate to compare the changes in adipose tissue mass (both total and in the lower limbs) during the intervention period as compared to the control period. Model assessment using various types of residual analyses may lead to performing the LMM on a transformed scale. | | reduces adipocyte mass in patients.<br>If no significant difference is found ($p < 0.05$), we will interpret this as we have no evidence that regular exercise affects adipocyte mass. | |
| 3. Determine whether 12 weeks of HIIT improves health-related quality of life in women with lipedema. | H1: 12 weeks of HIIT improves health-related quality of life in lipedema patients.<br>H0: 12 weeks of HIIT has no effect on health-related quality of life in lipedema patients. | N= 58, the same sample detailed above will be used to address this question. | We will compare changes in self-reported quality of life in the intervention period and the control period using the SF36 questionnaire.<br>We will apply LMM with repeated measures (random | The same sample detailed above are used. | If we find a significant difference ($p < 0.05$), we will interpret this as evidence that regular exercise improves quality of life in patients.<br>If no significant difference is found | N/A |
|  |  |  | intercept model) using age and baseline adipose tissue mass (%) in the lower limbs as covariates to compare the changes in quality of life during the intervention period as compared to the control period. Model assessment using various types of residual analyses may lead to performing the LMM on a transformed scale. |  | ($p < 0.05$), we will interpret this as we have no evidence that regular exercise improves quality of life. |  |

HIIT, high intensity interval training; BPI, Brief Pain Inventory; LMM, linear mixed models.

### Primary aim

Determine whether 12 weeks of HIIT improve pain levels in women with lipedema.

### Secondary aims

Determine whether 12 weeks of HIIT affects the following parameters:

I. reduces total adipose tissue mass and adipose tissue mass in the lower limbs.
II. improves health-related quality of life.

### Hypotheses

Our **1**^**st**^ **hypothesis** is that 12 weeks of HIIT will improve pain levels in women with lipedema. Our **2**^**nd**^ **hypothesis** is that the HIIT will I) alter body composition by decreasing adipose tissue mass, especially in the lower limbs, and II) improve quality of life.

### Goals and deliverables

The short-term goal is to establish the first evidence of the effects of HIIT in women with lipedema. The long-term goal is to improve the guidelines for treatment strategies of lipedema. The main deliverables from LipidEx includes increased knowledge of the effects of HIIT on pain, adipose tissue, body composition, and quality of life in women with lipedema. The findings may have significant value for lipedema management and potential to change clinical practice. By investigating the effects of HIIT in both the physiological and psychological aspects of lipedema, this project may provide a valuable foundation for the development of evidence-based, patient-centered treatment approaches.

## Methods

### Ethics information

The study will be approved by the Regional Committee for Medical Research Ethics (795836) and will be performed in line with the Declaration of Helsinki and Good Clinical Practice. Data Protection Impact Assessment (DPIA) will be performed and approved by the Department of Circulation and Medical Imaging at the Norwegian University of Science and Technology (NTNU). Informed consent will be obtained from all participants. The study is registered at clinicaltrials.gov (NCT06558851). The participants will be able to reserve themselves from tissue biopsies and still participate in the study.

### Design

This study will follow the Standard Protocol Items: Recommendations for Interventional Trials (SPIRIT) reporting guidelines.^45^

### Study design and participants

We will conduct a randomized controlled crossover trial (RCT) with 58 women diagnosed with lipedema. To be eligible for participation, women must be between 18 and 65 years old and have a confirmed diagnosis of lipedema. Exclusion criteria include self-reported ongoing eating disorders, pregnancy, regular endurance training, medications for weight loss, and/or orthopedic limitations for exercise training. Eligible participants will be randomized (1:1) using block randomization in eFORSK, an online randomization tool for clinical trials, to either Training group 1 or Training group 2. Training group 1 will start with 12 weeks of HIIT, tailored to each participant’s fitness level and the severity of their lipedema, and Training group 2 will serve as a control group during this 12-week period by continuing their usual lifestyle. Following the 12-weeks of intervention, Training group 1 will proceed into a 12-week wash-out period followed by a 12-week control period where they resume to their usual lifestyle. Training Group 2 will start their 12-week HIIT intervention period after their control period. Each participant will attend 3 or 4 test days, depending on the allocated group, where they undergo different measurements and testing. The study duration will be approximately 24 or 36 weeks.

### Recruitment

Participants will be recruited in Trøndelag, Norway, primarily through The Clinic of Rehabilitation at St. Olavs Hospital and the Norwegian Lymphedema and Lipedema Foundation (NLLF). Recruitment posters will be displayed at general practitioner offices and various physiotherapy clinics in the Trondheim area and spread thorough the social media of NLLF and the internationally renowned Cardiac Exercise Research Group (CERG). Interested participants can register via a QR code on the poster and will be contacted shortly thereafter to determine eligibility. Subsequently, they will receive detailed information about the study to evaluate their interest in participation.

### Exercise intervention

Exercise testing and training will be conducted at state-of-the-art exercise training facility at the Norwegian University of Science and Technology (NTNU), the *NextMove* Core facility, supported by highly experienced staff. The participants will perform three sessions of HIIT per week for 12 weeks. HIIT involves 4 times 4-minutes intervals at 85-95% of maximal heart rate (HR_max_) with 3-minutes active breaks (∼60% HR_max_) between the intervals. Two HIIT sessions are supervised at the *NextMove* core facility, and performed on treadmills or spinning bikes, while the last session of HIIT is unsupervised. In this unsupervised session, swimming will be recommended based on the potential benefits gained from compression. The criteria for adequate compliance to the exercise intervention program will be set to ≥80%. With standardized templated from the *NextMove* core facility, the participants are required to write a diary of their physical activity during the entire study period, including the exercise intervention period, the wash-out period, and the control period. All participants will receive a heart rate monitor to use throughout the intervention period. After the study is completed, the participants are welcome to keep the heart rate monitor, with the intention to promote a healthy lifestyle following the study.

### Data collection

During the first study visit, participants will be randomized into groups and receive comprehensive information about the study and all included procedures. Data collection will occur primarily on three or four separate test days, depending on the Training group, with a duration of approximately 3 hours. On these test days, anthropometric and physiological measurements (blood pressure and maximal oxygen uptake (VO_2max_)) will be taken, questionnaires will be completed, and fasting blood samples will be collected. Adipose tissue samples will be collected for those who consent, on the test day before and after the intervention period (two samples collected per participant in total). General information, including details on lipedema (type and morphological stage, and time of diagnosis), comorbidities, sleep patterns, and the use of medications, supplements, and clinical aids (e.g., compression aids and their frequency of use), will also be gathered on all test days. During the intervention period, the participants will be asked to fill out a short questionnaire at their last training session each week to gather information about perceived pain and health-related quality of life. All data from the test days will be registered in eFORSK.

### Pain registration

Pain will be registered at all test days using the RAND-36 health survey (Pain Dimension) (**primary endpoint**).^46^ RAND-36 (also known as SF-36) includes two questions designed to measure the intensity of pain and the extent to which pain interferes with normal work (both outside the home and housework). The first question assesses the intensity of pain experienced over the past four weeks with six response options from “none” to “very severe”. The second question evaluates the degree to which pain has interfered with daily activities and work with five response options from “not at all” to “extremely”. In addition, pain will be registered before and after both the intervention period and the control period using the questionnaire Brief Pain Inventory (BPI)^40^. This questionnaire includes four questions that assess the severity of pain right now and at its worst, least, and average. Each question gives a possible answer of 0 to 10, were 0 means no pain and 10 is the worst pain you can imagine. The four BPI questions contribute with the same weight, and they are combined to a final pain severity score with a range from 0 to 40. The results from RAND-36 and BPI will be evaluated separately. Whereas both tools assess pain, the BPI provides a deeper and more detailed assessment of pain characteristics and functional impact compared to the broader, more generalized assessment provided by the RAND-36 pain dimension. However, the questionnaires focus on different timeframes, as RAND-36 assesses pain over a retrospective period of the past 4 weeks whereas BPI assess pain over the past 24 hours or at the present moment (current pain intensity). Potentially, RAND-36 can be more useful for assessing chronic pain, and BPI for assessing more current pain intensity. However, the most suitable tool for assessing pain specifically in lipedema has yet to be determined.

### Anthropometric measurements

Body composition, including weight (kg), body mass index (kg/m^2^), muscle mass (kg), adipose tissue mass (%), basal metabolic rate (kcal), and mineral status (kg), will be measured non-invasively using the bioimpedance instrument InBody 770 (Biospace CO, Ltd, Seul, Korea). The instrument provides total adipocyte mass and adipose tissue mass in the lower limbs (**secondary endpoints**). The heigh (m) will be measured without shoes. The circumference (cm) of the waist, hip, right calf, and right thigh will be measured with standard procedures at *NextMove*, and the waist-to-hip ratio (cm/cm) will be calculated. A measuring board will be used to ensure similar site for pre- and post-measurement of the right calf and thigh.

### Quality of life

The RAND-36 questionnaire will be used to detect potential changes in health-related quality of life and data will be collected at all test days (**secondary endpoint**).^46^ This questionnaire includes 36 questions covering eight health domains, including vitality, physical functioning, bodily pain, general health perceptions, physical role functioning, emotional role functioning, social role functioning, and mental health. Each domain is scored on a scale from 0 to 100, where the scaled scores are weighted sums of all questions within each of the eight domains. A higher score indicates better health and well-being.

### Physiological measurements

Blood pressure will be measured in a sitting position after a minimum of five minutes rest in a quiet room with a handheld sphygmomanometer (Tycos, 5098-02CB, USA) by trained personnel. The blood pressure will be measured at the same time-point of the day for each participant at all test days. The first reading will be discarded and the mean of the next three consecutive readings will be used. Additional readings will be required if the coefficient of variation is above 15%.

VO_2max_ will be measured using ergospirometry (Metalyzer ll, Cortex Biophysik GmBH, Leipzig, German) supervised by qualified personnel. To demonstrate cardiometabolic effects from the HIIT intervention, VO_2max_ will be measured during uphill treadmill walking or running. A 10-minute warm-up (∼60% of HR_max_) will be performed before increasing the workload (1 km/h in speed or 2% in incline) every minute until exhaustion. The criteria for reaching VO_2max_ is a levelling off oxygen uptake (VO_2_) despite increased workload, and a respiratory exchange ratio of ≥1.05. Heart rate will be measured continuously during the test with Polar belt and watch (Polar, Polar Electro, Kempele, Finland), and the data will be used to determine HR_max_.

### Blood samples and adipose tissue biopsies

Four fasting venous blood samples will be drawn by qualified personnel and by standard in-hospital procedures at St.Olavs Hospital. Directly after collection, one 3mL EDTA-tube and one 3mL Li-heparin-tube will be sent to routine biochemical analyses at St. Olavs hospital for measuring glucose, HbA1C, triglycerides, HDL-cholesterol, total cholesterol, lipoprotein (a), serum ferritin and hs-CRP. The two remaining tubes (serum and EDTA) will be centrifuged, aliquoted, and stored in the LipidEx biobank at -80°C for explorative analyses.

Two adipose tissue biopsies will be collected by experienced personnel from the Research outpatient clinic at St.Olavs Hospital before and after the exercise intervention. The collected adipose biopsies will be used to advance our physiological understanding of exercise-induced effects on adipose tissue in women with lipedema. To ensure patient comfort during the procedure, local anesthesia will be administered at the site of biopsy to minimize pain and discomfort associated with the needle insertion. The proposed methodology will adhere to rigorous sterilization protocols, including the use of sterile equipment, gloves, and disinfection procedures. A fine needle is used to avoid the need for extensive incisions and to reduce patient discomfort and recovery time. One biopsy will be snap-frozen in liquid nitrogen and stored at - 80°C for later explorative analyses. Another biopsy will be placed in histology cassettes and fixed in 4% formaldehyde in 0.1 M phosphate buffer for 24 hours. The samples will be dehydrated, embedded in paraffin, sectioned, and stained with hematoxylin and eosin at the general lab at the Institute for Circulation and Medical Imaging, NTNU in later explorative analyses.

## Explorative analyses

These analyzes will be reported under the heading explorative analyses if the protocol proceeds to Stage 2.

## Sampling plan

### Statistical power

As lipedema is a disease that just recently has received more attention in research, there is limited data available in the literature to make an accurate prediction of the expected size of the effect of HIIT on pain levels in women with lipedema. Thus, for calculating the number of participants needed to obtain a power of 80% at a significance level of 0.05, we used data from our recent feasibility study (ongoing trial, ClinicalTrials.gov NCT05488977). 22 participants have so far completed the feasibility study, and we used the collected data and observations from these participants. The mean self-reported pain level at baseline for the lipedema patients were 55 points (based on RAND-36), and the standard deviation was 23. We consider a reduction in pain levels of 10 points to be clinically relevant, which equals a change to 65 points (inversed pain scale). Based on this, a sample size of 44 participants would be included in the study to achieve a power of 80% at a significance level of 0.05 (assuming a correlation of 0.5 between pre- and post-measures of pain). When accounting for up to 30% drop out (due to the length of the trial), we would need to include 58 participants.

## Analysis Plan

### Pre-processing steps

Data is collected by the electronic case report form (eCRF) eFORSK. After data cleaning (searching for data entry errors or outliers), the database will be locked, and data will be extracted as sav files and csv files for SPSS and R, respectively. The following statistical analyses will be performed using SPSS v.21 and RStudio.

Efficacy of the intervention will be assessed on an intend-to-treat approach, where all trial data will be analyzed regardless of compliance with the intervention. In addition, per-protocol analyses will be conducted where we include participants who have completed at least 80% of the planned exercise training sessions (including both the supervised and non-supervised sessions). Interim analyses are not planned. Sample description will be done via frequencies and corresponding percentages, medians (with interquartile range), or means (with standard deviation).

### Planned analyses

### Pain – Primary endpoint

We will apply linear mixed models (LMM) with repeated measures (random intercept model) using age and baseline adipose tissue mass (%) in the lower limbs as covariates to compare the changes in pain levels during the intervention period as compared to the control period. Model assessment using various types of residual analyses may lead to performing the LMM on a transformed scale.

### Adipose tissue mass (%) – Secondary endpoint

We will apply LMM with repeated measures (random intercept model) using age as covariate to compare the changes in adipose tissue mass (both total and in the lower limbs) during the intervention period as compared to the control period. Model assessment using various types of residual analyses may lead to performing the LMM on a transformed scale.

### Quality of life – Secondary endpoint

We will apply LMM with repeated measures (random intercept model) using age and baseline adipose tissue mass (%) in the lower limbs as covariates to compare the changes in quality of life during the intervention period as compared to the control period. Model assessment using various types of residual analyses may lead to performing the LMM on a transformed scale.

### Contingency Analysis

### Pain Reduction Impact on Secondary Outcomes

If a significant reduction in pain is observed, we will perform mediation analysis using appropriate models, such as ordinal generalized linear mixed models (GLMMs), to explore whether changes in secondary outcomes (total adipocyte mass (%), adipocyte mass in lower limbs (%), or quality of life) mediate the relationship between HIIT and pain reduction.

### Responder analysis

Based on the results we will identify responders and non-responders to HIIT based on pain reduction. Those who achieved a 5-point reduction in pain levels after HIIT will be assigned as responders, and those with no reduction or even an increase in pain levels during the HIIT will be assigned as non-responder. Those with 0-5-point reduction in pain levels will be excluded from responder analysis. Then we will compare secondary outcomes between responders and non-responders using t-tests or Mann-Whitney U tests if normality assumptions are not met.

## Data availability

We commit to share raw data and materials on acceptance of the Stage 2 manuscript.

## Code availability

We commit to share all codes on acceptance of the Stage 2 manuscript. The codes will be made open access at this location: https://github.com/anjabye/LipidEx.

## Acknowledgements

The study was supported by grants from the DAM Foundation and Joint Research Committee between St. Olavs Hospital and the Faculty of Medicine and Health Sciences, NTNU (FFU). The funders had/have no role in the study design, data collection and analysis, decision to publish, or preparation of the manuscript.

## Author contributions

JCS: Conceptualization, supervision, methodology, project administration, writing – original draft

RMP: Conceptualization, methodology, writing– review & editing

RU: Writing– review & editing

LM: Conceptualization, methodology, writing – review & editing

ILN: Methodology, writing – review & editing

SK: Conceptualization, writing review & editing

KMO: Methodology, writing – review & editing GE: Writing – review & editing

TB: Writing – review & editing JL: Writing – review & editing

SN: Conceptualization, methodology, project administration, writing – original draft

GFG: Conceptualization, methodology, writing – original draft

TF: Methodology, writing – original draft

AET: Conceptualization, methodology, project administration, writing – original draft

AB: Conceptualization, supervision, methodology, project administration, writing – original draft

